# Search for the trend of COVID-19 infection following Farr’s law, IDEA model and power law

**DOI:** 10.1101/2020.05.04.20090233

**Authors:** Srijit Bhattacharya, Moinul Islam, Alokkumar De

## Abstract

Following power law, Farr’s law and IDEA model, we analyze the data of COVID-19 pandemic for India up to 2 May, 2020 and for Germany, France, Italy, the USA, Singapore, China and Denmark up to 26 April, 2020. The cumulative total number of infected persons as a function of elapsed time has been fitted with power law to find the scaling exponent (*γ*). The reduction in *γ* in different countries signals the reduction in the growth of infection, possibly, due to long-term Government intervention. The extent of infection and reproduction rate *R*_0_ of the same are also examined using Farr’s law and IDEA model. The new cases per day with time assume Gaussian bell shaped curve, obeying the rule that faster rise follows faster decay. In India and Singapore, the peak of the bell shaped curve is still elusive. It is found that, till date, countries such as Denmark and India implementing sooner lockdown have underwent lower number of new cases of infection. Daily variation shows, *R*_0_ of all the countries is reducing, ushering in fresh hopes to combat COVID-19. Finally, we try to make a prediction as to the date on which the different countries will come down to daily cases of infection as low as one hundred (100).

## I. INTRODUCTION

The outbreak of communicable infectious disease is a major threat to the human civilization. The present novel corona virus (COVID-19) pandemic corroborates this, albeit the humongous scientific advancement in the fields of medicines and vaccinations. As of 28th April, 2020 11:59 pm (Indian Standard Time), the total number of confirmed COVID-19 case is overwhelmingly large, about 3060152 globally. Out of the infected persons, the death toll is about 7% (211894 persons) [1]. Thus the very pandemic elaborates the need for timely identification and proper surveillance of this infectious disease. Towards this aim, mathematical modelling might be a key. Through the modelling, by predicting the outcome, the dynamics of spread can be understood and correspondingly the future course of action can be planned. The Royal Society Committee [2, 3] on infectious diseases affirmed that ‘Quantitative modelling is one of the essential tools both for developing strategies in preparation for an outbreak and for predicting and evaluating the effectiveness of control policies during an outbreak…More work is required to refine the existing models and to strengthen their capacity to inform policy.’ However, the success of data-driven predictive models depends a lot on the reliability of the available datasets. Generally, SIR (susceptible infected recovered) has been most commonly used mathematical model to identify disease outbreak and its trajectory [4]. It is also used to predict COVID-19 pandemic. Besides, stochastic models including Markov-chain based algorithms [5] have also been used in predicting different disease outbreaks. In addition to such rigorous mathematical models, simple mathematical formula based models have also been found successful to predict the future course of infection. Power law formula is one of them. Starting from as abstract as frequency of words in English poems, the income distribution, short term human travel behavior, city size distribution, etc. have been well-explained by power law formula [6, 7]. It is also used in predicting the trend of different infectious diseases [8]. Recently [9], the trajectory of COVID-19 disease growth has been described by power law formula. In spite of the over-prediction done by power law in the outbreak of Severe Acute Respiratory Syndrome (SARS) and Middle East Respiratory Syndrome (MERS) diseases, the success of this law in COVID-19 prediction is notable [9].

With reference to COVID-19, we explained in a recent work [10], the nature of variation of the cumulative number of observed infected persons *n*(*t*) as a function of elapsed time for different countries for a time span from 22 January, 2020 to 1st April, 2020 in terms of power law *n*(*t*) = *At*^*γ*^, where *γ* is power law exponent and *A* is constant. The value of *γ* is related to the rapidity of the spread of disease in a country. We found that, up to 1st April, the infection moved to a saturation (*γ* is reduced from 2.18 to nearly zero i.e 0.05) in case of China, whereas in Denmark the infection was in slow down stage (*γ* reduced from 6.82 to 1.48). We calculated the change in *γ* from the change in slope of the power law graph (in log-log scale). We also noticed that the extent of outbreak could not be demonstrated completely via this law. We could only compare the infection trajectory between different countries and inferred whether the rate of infection was gradually reducing in a country from the value of the exponent in comparison to others. Moreover, in the existing literature [8], the failure of power law model in the identification of disease outbreak has been seen. Therefore, modification of power law has always been sought for.

For proper surveillance of an infectious disease, identification of start and end of the same is essential along with the understanding of its strength. Farr’s theory of epidemics, or Farr’s law in short, states that epidemics rise and fall with time in the symmetrical pattern, producing a bell shaped curve [11]. The law demonstrates the growth and decay periods of an infectious disease. However, to understand the strength of an infectious disease, it is highly important to estimate the reproduction number *R*_0_ of the infection. *R*_0_ *<* 1 suggests each existing infection is producing less than one new secondary infection, which means the disease will decay and ultimately die out. On the contrary, *R*_0_ > 1 means an epidemic with each infection causing more than one secondary infection. The value of *R*_0_ is implicitly embedded within the Farr’s law [12–14]. The two-parameter Incidence Decay with Exponential Adjustment (IDEA) model [14], on the other hand, can estimate the reproduction number of infection explicitly. IDEA model also assumes that the peak and decay of infectious disease are not solely caused by the depletion of patients. The measures taken by authority, the change in behaviour by general public, etc. all play important roles to control the outbreak. The gross result is reflected in the *R*_0_ value. In this work we use Farr’s law and IDEA model, both, to explore the strength of COVID-19 outbreak in India, the United States of America (USA), Italy, Germany, France, China, Singapore and Denmark. We utilize Farr’s law to identify the start and end of infection in terms of symmetric Gaussian distribution. The power law fit is also exploited to gather knowledge in the relative change in cumulative number of infection. Finally, the prediction has been made as to probable future situation.

## II. METHODS

The daily data of infected persons have been extracted from JHU-CSSE 2020 open data source of John Hopkins University [1]. For India, the dataset is extracted from the Website of Ministry of Health and Family Welfare, Govt. of India [15]. The cumulative number of infected person (*n*(*t*)) is plotted against elapsed time. The power law fit is done in log-log scale and the least square best fit value of *γ* is extracted, as explained in ref. [10].

### 1. Farr’s law

Farr’s law relates, empirically, the number of new cases of observed infected persons within successive time intervals in an epidemic by the following relation:

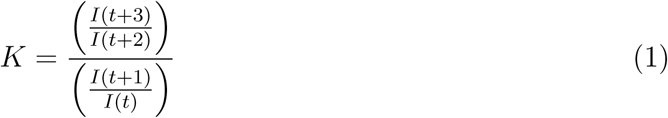

where *I*(*t*) is the number at time *t* and *K* is constant.

For *K* < 1, the rate of change of observed new cases decreases with time. *I*(*t*) satisfying Eq. 1 follows the trend of bell-shaped Gaussian or normal distribution [14, 16]. We fit the bell shaped part of the graph of *I*(*t*) against elapsed time (*t*) by the following Gaussian function:

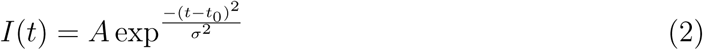

where *A* is normalization constant, *t*_0_ is centroid of the Gaussian curve and *σ* is related with the standard deviation of the distribution. Here elapsed time *t* is expressed in days.

The pattern of smallpox deaths in England and Wales during 1837-39 closely resembled the trend as governed by the Eq. (2). John Brownlee [12] observed that *I*(*t*) followed the function, exp (−*At*^2^ + *Bt* + *C*), where, *A*, *B* and *C* are constants and the resultant curve is bell shaped, similar to the result of Farr’s law. Thus Brownlee identified that an epidemic would be related to the growth of new cases (a first order process) and the decay (second order). This is analogous to IDEA model.

### 2. IDEA model

IDEA model, in a single equation, describes an epidemic process with first order exponential growth and second order simultaneous decay. The growth has been a function of *R*_0_[13, 14]. The temporal evolution of infected persons satisfies the following equation:

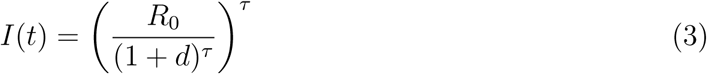

where *τ* is generation time from primary to secondary infection. In case of COVID-19 infection, this incubation is usually of 14 days period [16]. So *τ* =1, 2, 3, …, 14. *d* (here *d* > 0) is a ‘control’ parameter that reduces the new cases of infection. Control comes in different forms like lockdown, social distancing, health measures taken by Government, growth of immunity within people, availability of personal protective kits, vaccinations or medications, etc. The IDEA model and Farr’s law can be inter-related by the relation, *K* = 1/(1 + *d*)^4^. IDEA model demonstrates that when *I*(*t*) is expressed as exp (−*At*^2^ + *Bt* + *C*), the constants *A* and *B* become log (1 + *d*) and log *R*_0_, respectively. Thus, comparing Eq. 2 and exp (−*At*^2^ + *Bt* + *C*), we calculate *A* and *B*. Then the values of *d* and *R*_0_ are estimated from *A* and *B*.

## III. RESULTS

### A. Power law

In figs. 1 and 2, the cumulative number of observed infected persons have been plotted against elapsed time (in day) in log-log scale and fitted with power law, for the countries viz. the USA, Denmark, China, Italy, India, Germany, France and Singapore. As per the figures, the rate of infected persons has been decreasing in the countries studied, except for Singapore, after the Government policy interventions. This should be reflected in the *γ*-values. Thus, the initial and preceding *γ*-values, have been termed as pre (*γ_pre_*) and post interventions (*γ_post_*), respectively [10, 17]. The power law fit of the observed data of Denmark (fig. 1 panel B) gives pre-intervention *γ* or *γ_pre_*=6.82. The lockdown effect reduces *γ* from 6.82 to 1.48 (the dataset showing pre-intervention is represented by red filled circles while the *γ_post_* is shown by the datasets with open circles in the same figure). The post-intervention *γ* again reduces to 1.23 (shown by open triangles). Similarly, in fig. 2 panel C, for France *γ* is reduced from 10.81 to 4.65, and presently to 0.88 only. Panel D of the same figure is the trend in Singapore, where *γ* has not been reduced from the initial value of 10.80. More than one slope is evident in the slowing down or post-intervention stage only in Denmark, Germany and France. The saturation in cumulative data could not at all be seen in the countries studied, except in China. Indication of saturation is found in France, but more data are needed for conclusive evidence. Table I gives the country wise values of *γ*.

**FIG. 1.**
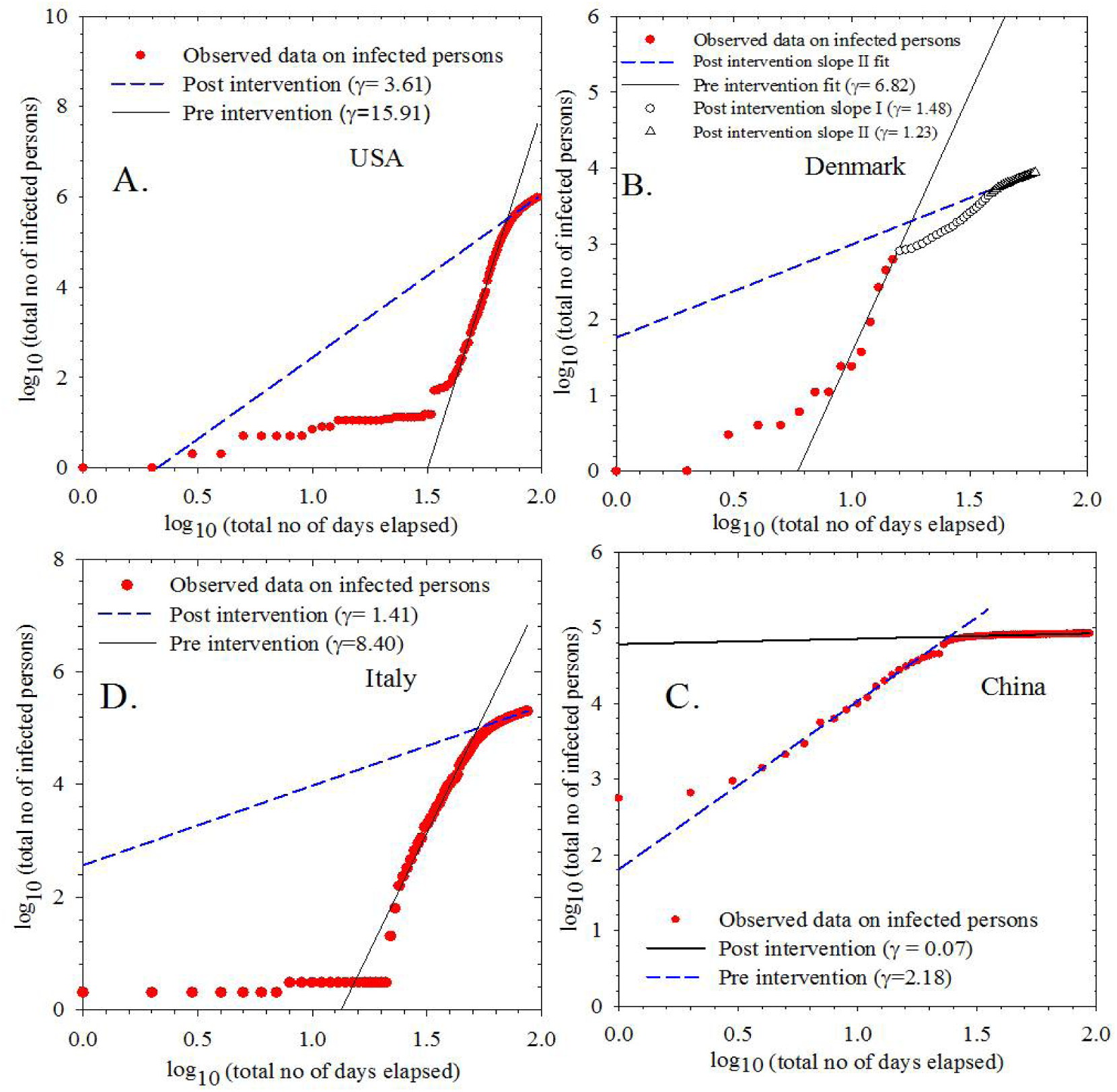
(Online colour) COVID-19 infection in the USA, Denmark, China and Italy (in the panels A to D, clockwise from top left), plotting being done in log-log scale. Red filled circles are the observed data points. In Denmark, the open circles and triangles are data points having reduced slopes. The two straight lines (blue dashed and black solid) are the yields of linear regression for pre-intervention and post-intervention, respectively. The starting dates of the datasets are 22 January, 27 February, 22 January and 31 January, 2020, respectively, corresponding to the panels A to D.

**FIG. 2.**
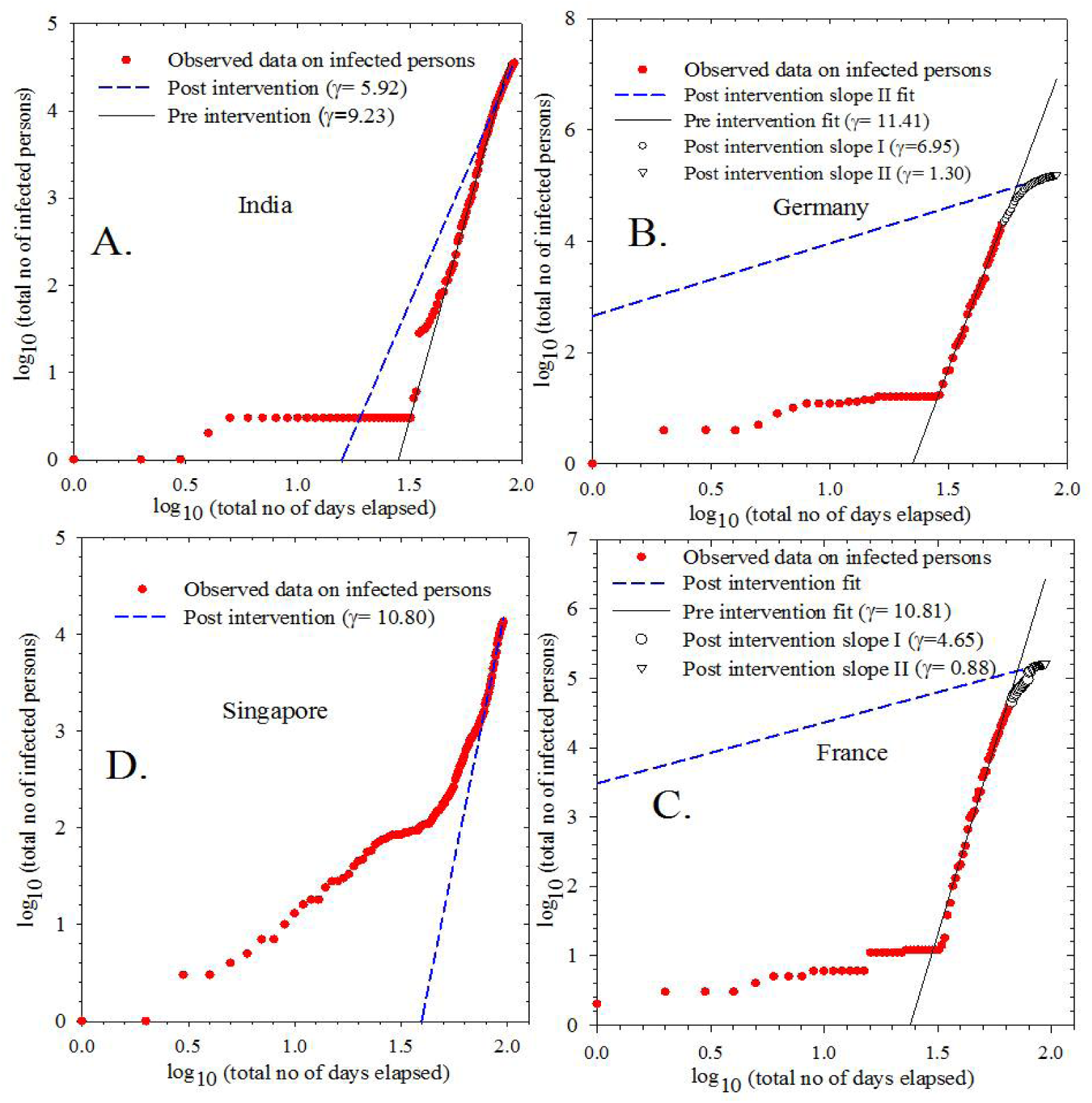
(Online colour) COVID-19 infection in India, Germany, France and Singapore (in the panels A to D, clockwise from top left), plotting being done in log-log scale. Red filled circles are the observed data points. In Germany and France, the open circles and triangles are data points having reduced slopes. The two straight lines (blue dashed and black solid) are the yields of linear regression for pre-intervention and post-intervention, respectively. The starting dates of the concerned datasets are 30 January, 26 January, 24 January and 22 January, 2020, respectively, corresponding to the panels A to D.

**TABLE I.**
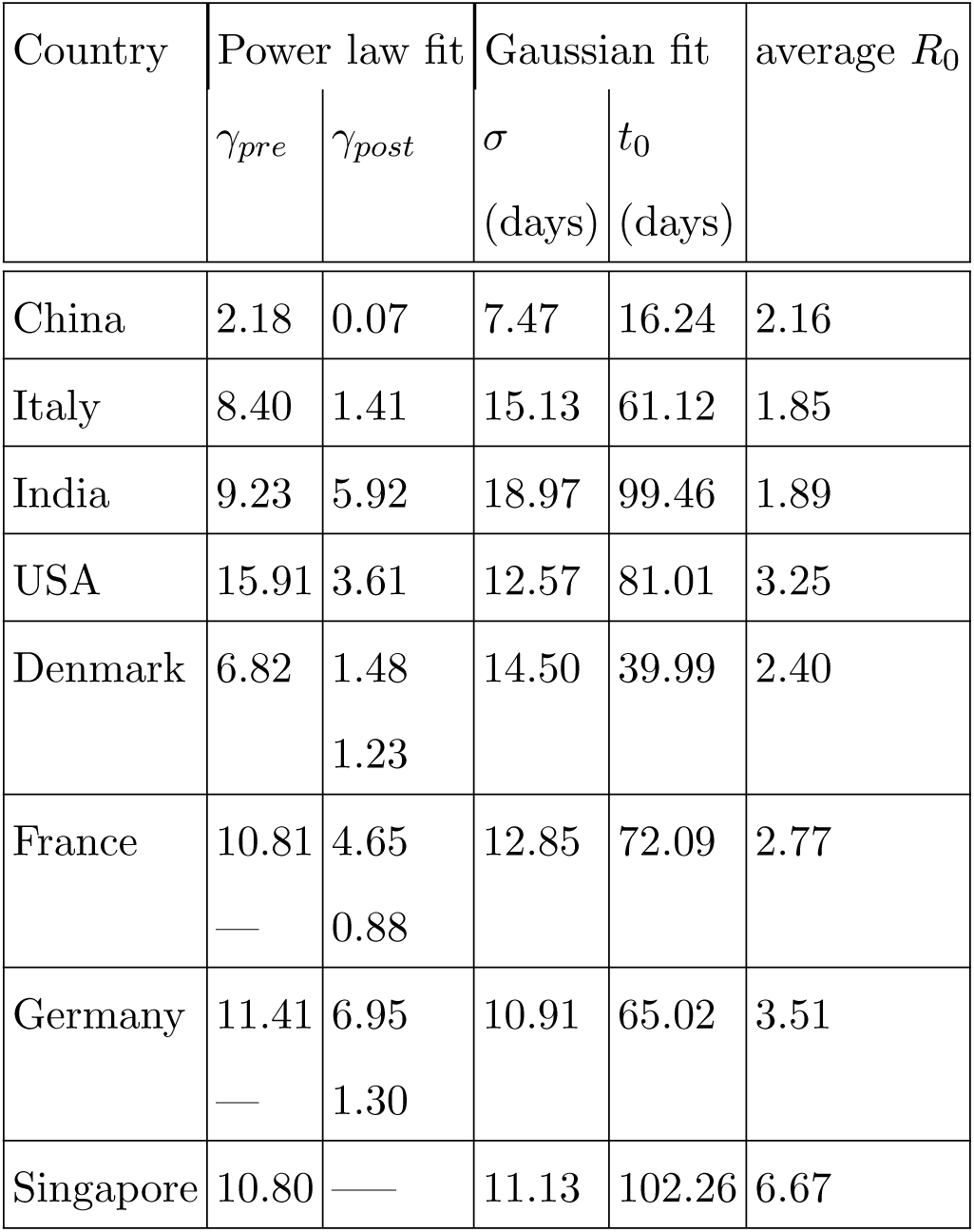
The list of power law exponents *γ_pre_*, *γ_post_*, standard deviation (*σ*), centroid (*t*_0_) of the Gaussian curve and reproduction rate *R*_0_ for different countries.

### B. Farr’s law

The new cases of infection per day is plotted against the time elapsed in terms of date and shown in figs. 3 and 4. The datasets of all the countries show symmetrical bell shaped nature with gradual rise and decay. The bell shaped part is fitted with Gaussian function as in Eq. (2). The best fitted values (deduced from *χ*^2^ minimization technique) of the standard deviation (*σ*) and centroid (*t*_0_) of the Gaussian function are mentioned along with the graphs for each countries. For, India, Singapore and the USA, 97% of the observed data lie within ±1*σ*. For Italy, Germany and France, about 75% of the observed data lie within ±1*σ* and 96% within ±2*σ*. For China and Denmark, the fluctuation in daily data increases the error of the fit. Only about 50% of the data are within ±1*σ* and 75% are within ±2*σ*. Skewness of all the distributions is estimated and found within 0.4 to 0.6. Kurtosis is also calculated and found within −1.2 to −1.4. These values indicate symmetric normal distributions for all the cases studied here.

**FIG. 3.**
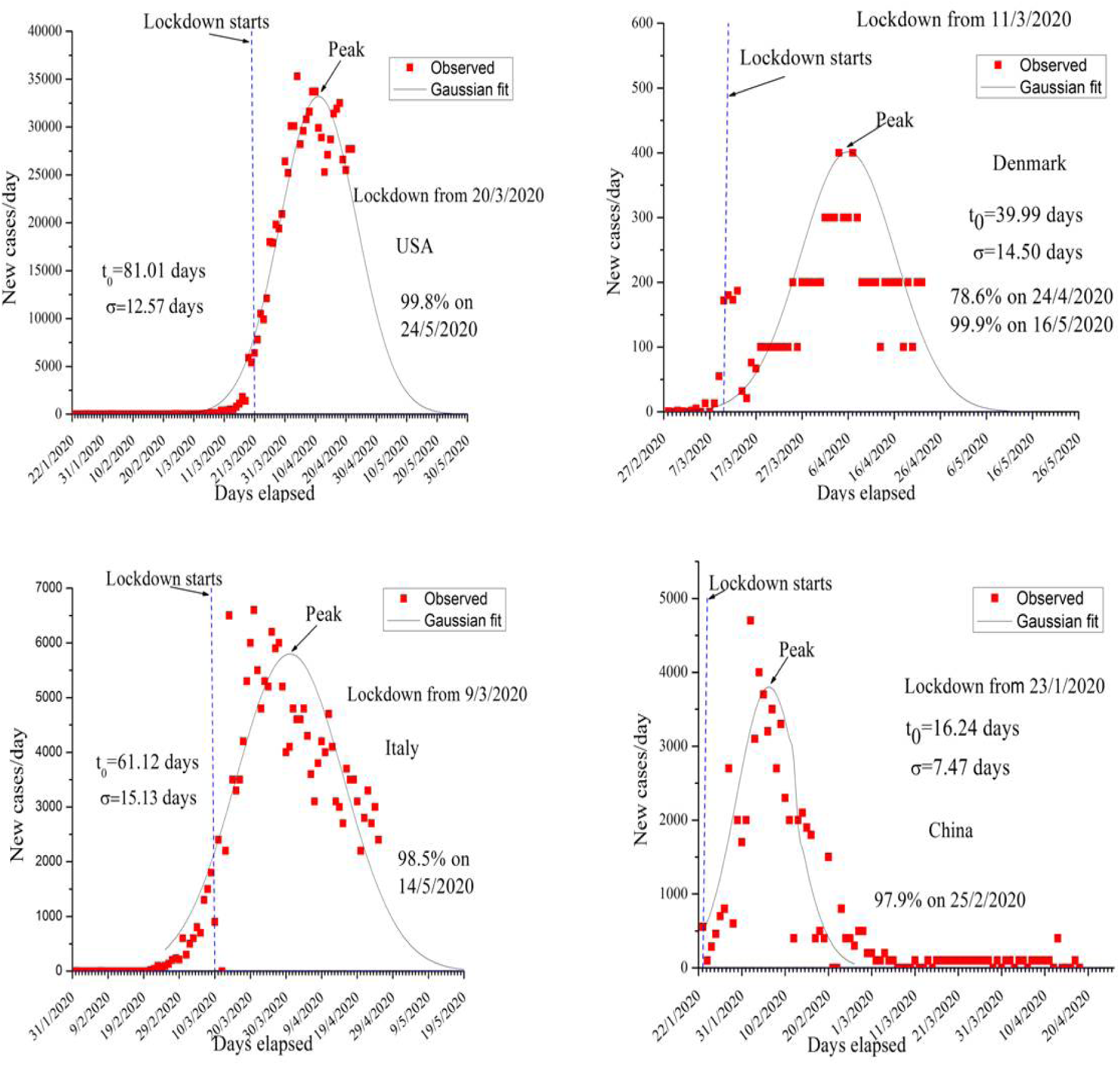
(Online colour) Farr’s law validation in the new COVID-19 infection per day with elapsed date for the countries the USA, Denmark, China and Italy (clockwise from top left). Red filled circles are the observed data points and black continuous line is the Gaussian fit. The concerned date for decay below 100 infections per day and corresponding percentage are also shown in each plot.

**FIG. 4.**
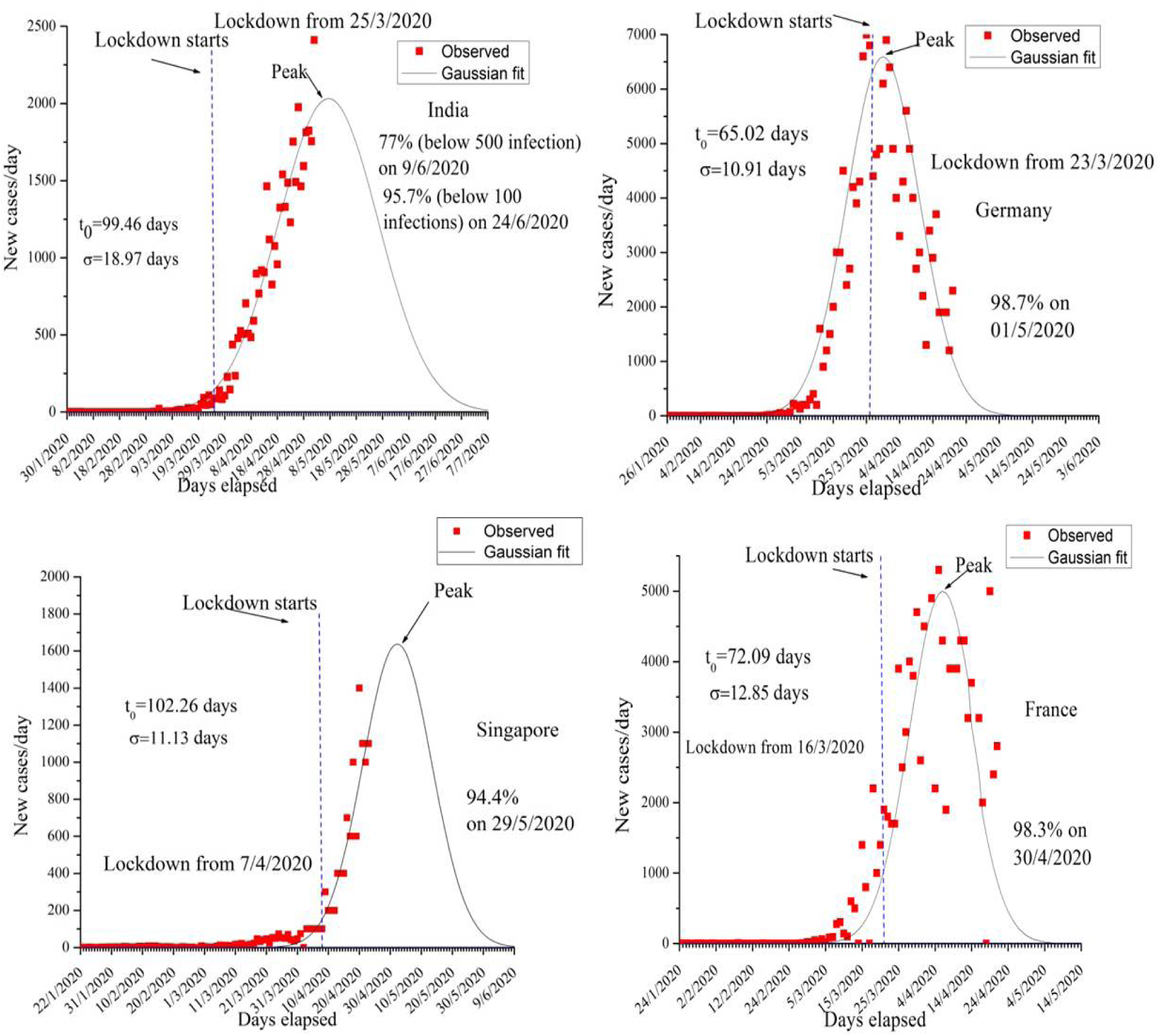
(Online colour) Farr’s law validation in the new COVID-19 infection per day with elapsed date for the countries India, Germany, France and Singapore (clockwise from top left). Red filled circles are the observed data points and black continuous line is the Gaussian fit. The date and percentage of corresponding decay below 100 infections per day are shown in each plot. For India, the date and percentage of corresponding decay below 500 infections per day are also mentioned.

### C. IDEA model

Using the expression exp (−*At*^2^ + *Bt* + *C*) and Eq. 3, the time dependent values of *R*_0_ and d are calculated. The value of *R*_0_ has been plotted against elapsed time in fig. 5 and compared for three different countries (India, the USA and Italy). The average values of *R*_0_ are explored for all the countries studied here. Table I lists the countrywise values of centroid (*t*_0_), standard deviation (*σ*) and *R*_0_.

**FIG. 5.**
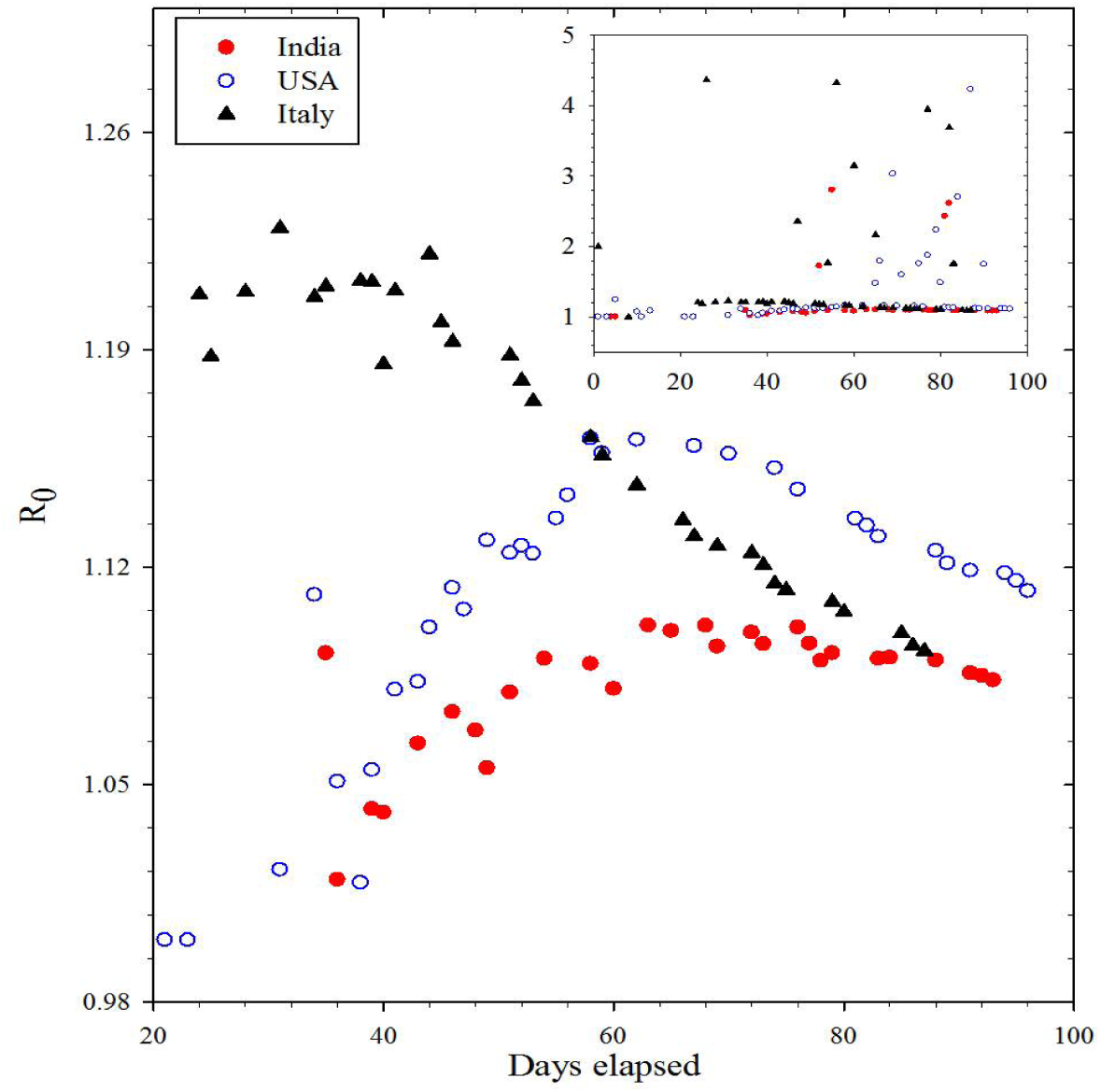
(Online colour) Plot of *R*_0_ against number of days elapsed. The plots are also compared for India, the USA and Italy. Inset shows the same graph with different scaling to understand the complete variation of *R*_0_.

## IV. DISCUSSIONS

From the power law fits, it is clear that except in Singapore, presently the rate of infection has decreased in the other countries. In China, the saturation in cumulative data arises from the 25th day of the starting date of the data (starting date is 22 January, 2020). Thus the value of *γ* becomes nearly zero (*γ*=0.07). Similarly, in France *γ* has achieved sub-decimal value of 0.88, though one order higher than that of China. Besides, in Germany and Denmark, the exponent has reduced significantly (by 89 and 82%, respectively, from the initial or ‘pre-intervention’ values). The USA, Italy and India show reduction in *γ* by nearly 77, 83 and 36%, respectively. Singapore has not yet shown reduction in *γ*. This decrease in *γ* has been explained in terms of Government intervention and different measures adopted. But the power law fit has not completely manifested the effect of Government intervention. Power law fit, in general, shows only the fractional rate change and may indicate the slowing down or saturation in the infection in progress [10].

In view of this, we have exploited Farr’s law to explore finer details of infection. Figs. 3 and 4 show the peaks of the normal curve along with corresponding dates. The Farr’s law points toward the fact that the new cases per day have still not achieved the peak in India and Singapore. This has been demonstrated by the lower reduction of *γ* in India in comparison to the USA or Italy. However, the extent of infection is much lower in India as shown by its peak value of the fitted Gaussian (approx. 2000 within first week of May, 2020) than in the USA (nearly 33K on 11 April, 2020) or Italy (nearly 5800 in 31 March, 2020). According to Farr’s law the rate of decrease in infection will follow the rate of increase. Our estimated full width at half maximum (FWHM) of the Gaussian curve also corroborates to this. The FWHM for India is very high (approx. 45 days) than that for the USA (approx. 28 days), Italy (approx. 34 days) and other countries studied here. Interestingly, if the countries are compared, it is found that as the lockdown date comes closer to the peak, the FWHM becomes smaller, which perhaps reiterates that the infection decay rate follows the growth rate. It is also observed that the earlier lockdown has given rise to lower peak values in different countries. In case of Germany, we found the FWHM of the Gaussian curve is only 24 days, whereas the lockdown date is closest to the peak (less than 1*σ*) than any other countries. This could imply the generation of immunity against COVID-19 infection in German people. But exception is found in China, where even after earliest implementation of lockdown and with smallest value of FWHM of the Gaussian curve, the maximum value of infection comes much higher than those for some other countries like India or Denmark.

From the IDEA model Eqs. 2 and 3, we have estimated daily variation of *R*_0_. From the value of the coefficient *B* in the expression *I*(*t*) = exp (*At*^2^ + *Bt* + *C*), the average value of *R*_0_ is also calculated. In the fig. 5, daily variation of *R*_0_ is compared among India, Italy and the USA. *R*_0_ has fallen from 1.95 on 18 April, 2020 to 1.08 on 31 April, 2020 in India. *R*_0_ was as high as nearly 5 in India on 23 March, 2020. According to a study done by Indian Council of Medical research (ICMR), India [18], the value of *R*_0_ was estimated as 4 on 23 March, 2020. It fell from 1.83 on 6 April, 2020 to 1.5 on 11 April, 2020. The values closely resembles to our study. Meanwhile in the USA, the value of *R*_0_ has been reduced from 1.74 to 1.11 upto 23 April, 2020. However, the value of average *R*_0_ is found highest in Singapore (6.67) and lowest (1.85) in Italy. The high value of *R*_0_ (2.40) in Denmark is possibly an artefact of large fluctuation in its daily data. For complete removal of COVIT-19, *R*_0_ should be less than 1. In spite of that, the reduction in daily variation of *R*_0_ indicates a glimmer of hope to combat COVIT-19.

Finally, we predict, from the Gaussian fitting, the date when the daily number of infection will be reduced to the value less than a hundred for all the countries. In the USA, around 99.8% of the infection will be reduced within 24 May, 2020. Italy will be 98.5% free from infection within 14 May, 2020, while India will be about 77% free from infection on 9 June, 2020. COVID-19 infection will come to an end by about 95.7% within 22 June, 2020 in India. The corresponding dates for other countries are mentioned in the figs. 3 and 4. Recent prediction done by Singapore University of Technology and Design (SUTD), using SIR model [19], is found in close agreement with our prediction for all the countries studied, except for India. In India, unlike SUTD prediction, we find that the very peak of the Gaussian distribution is elusive yet. However, before drawing any conclusion, it is to be noted that the analysis of dynamical data is always uncertain due to inherent volatility and fluctuation. These increase the uncertainty in prediction and may probably be reduced by continuous monitoring. Therefore, the mathematical model based prediction should be taken with proper caution.

It may be noted that in a few cases such as HIV disease in the USA, the performance of the prediction using Farr law was not satisfactory at all. Some of the reasons behind this could be the problem in datasets and lack of knowledge in the spread of a disease [20]. Therefore, we have investigated the spread of COVID-19 using three basic models such as power law, Farr’s law and IDEA model, on the same platform, to understand the spread of the disease as much as possible. Besides, SIR model prediction done by Singapore University of Technology and Design is also found in agreement with our study. Moreover, the spread of COVID-19 is completely different from that in HIV. This method, which is the combination of power law, Farr’s law and IDEA model, could be very strong yet simple framework to show the rate of infection, reproduction as well as the whole dynamics of infectious disease.

## V. CONCLUSION

The COVID-19 infection is fitted using power law scaling for the countries like the USA, China, Denmark, France, Germany, Italy and Singapore up to 26 April, 2020 and India up to 2 May, 2020. In continuation with our earlier work, we find that the value of γ reduces significantly in all countries, except in Singapore, where the post-intervention effects has not yet been seen. Using Farr’s law the new cases per day with elapsed time has been fitted with Gaussian function and decay of infection is predicted for the countries mentioned above. Combining IDEA model and Farr’s law, the reproduction rate *R*_0_ is also found. Interestingly, Farr’s law seems to show that countries with earlier lockdown usually suffer lesser number of infection. India and Denmark show smaller number of daily infections possibly due to earlier lockdown, though the peak in the Gaussian curve for India is still elusive. In terms of *R*_0_ and *γ*, it is clear that COVID-19 infection is gradually decreasing. The possible time required for the decay of infection is shown by the time in which the number of new cases becomes less than a hundred (100) per day. The whole technique adopted in this work appears to be a powerful yet simple one for the understanding of such infectious disease dynamics.

## Data Availability

All the data referred in our manuscript are available in the internet.

https://github.com/CSSEGISandData/COVID-19.

https://www.mohfw.gov.in/

https://coronavirus.jhu.edu/us-map

